# Protocol for a randomised, double-blind trial of a chronotherapeutic mobile health (mHealth) behaviour change intervention to optimise light exposure among older adults aged ≥60 years in Singapore (LightSPAN)

**DOI:** 10.1101/2025.09.12.25335635

**Authors:** Resshaya Roobini Murukesu, Zahrah Alwi Alkaff, Denz Del Villar, Johannes Zauner, Manuel Spitschan

**Affiliations:** TUMCREATE, Singapore, Singapore; TUM School of Medicine & Health, Department Health and Sport Sciences, Technical University of Munich, Munich, Germany; TUM Institute for Advanced Study (TUM-IAS), Technical University of Munich, Garching, Germany; Max Planck Institute for Biological Cybernetics, Max Planck Research Group Translational Sensory & Circadian Neuroscience, Tübingen, Germany

**Keywords:** digital health, older adults, chronobiology, behaviour change, light exposure

## Abstract

**Background:** Suboptimal light exposure among older adults can exacerbate circadian disruption, sleep disturbances, mood disorders, cognitive decline, and frailty. In urbanised environments like Singapore, older adults are particularly vulnerable due to lifestyle and built environment constraints. The LightSPAN study evaluates a chronotherapeutic mobile health (mHealth) behaviour change intervention, delivered via the LightUP app, designed to optimise light exposure patterns and support healthy ageing.

**Methods:** The main trial is a community-based, double-blind, parallel-group randomised controlled trial involving approximately 90 community-dwelling older adults (≥60 years) recruited through Active Ageing Centres in Singapore. Participants will be randomised using a secure web-based system with stratification by age, sex and recruitment site to receive either the active LightUP app with behaviour change features (goal setting, self-monitoring, personalised feedback) or a placebo version without these features. The primary outcome is daily time above 250 lx melanopic equivalent daylight illuminance (melanopic EDI) during daytime, measured with the ActLumus light logger across baseline, intervention and follow-up periods. Secondary outcomes include sleep parameters and circadian rest-activity rhythms, sleep quality, mood, cognition, frailty status, physical activity, body composition and vitamin D levels. The usability and acceptability of the LightUP app and wearable devices will also be assessed. Analyses will follow the intention-to-treat principle, supplemented by per-protocol analyses. Generalized linear mixed models will be used for repeated measures, with multiple imputation for missing data and exploratory Bayesian analyses. All analyses will be pre-registered on the Open Science Framework and de-identified datasets and code will be openly shared under a CC-BY license.

**Discussion:** Despite its central role in circadian regulation, light exposure has rarely been targeted in behavioural preventive interventions for older adults. This study evaluates a novel mHealth-enabled approach that empowers older adults to optimise their light exposure, extending the scope of non-pharmacological strategies for preventive medicine and health promotion.

**Trial Registration:** This study was retrospectively registered at the ISRCTN – The UK’s Clinical Study Registry on 05 September 2025 (ID: ISRCTN123919320, https://doi.org/10.1186/ISRCTN12391932).

## Background

Light is the dominant zeitgeber for human circadian rhythms, regulating sleep-wake timing, physiology and health^1^. Inadequate or poorly timed light exposure is associated with circadian disruption, impaired sleep, mood disturbances, cognitive decline and metabolic dysregulation^2–4^. Older adults are particularly vulnerable, as ageing is associated with reduced ocular light transmission, decreased outdoor activity and comorbidities that reduce daily light exposure^5–11^.

Evidence suggests that improving light exposure can stabilise circadian rhythms and support sleep and wellbeing in ageing populations, including patients with dementia^12–14^. However, conventional interventions such as bright light therapy boxes, while effective in controlled trials, often face barriers to real-world use: they require active engagement, can be costly and show low adherence^15^. Recent work has highlighted the need for scalable, accessible and user-friendly strategies that deliver sustained behavioural change in light exposure^16^.

Mobile health (mHealth) interventions offer a promising alternative. Smartphones can provide personalised feedback, encourage self-monitoring and deliver behaviour change techniques. While mHealth approaches have been widely studied for physical activity, diet and chronic disease management, little evidence exists on their role in modifying light exposure behaviour^16^ ^17^. Integrating wearable light sensors with mHealth interventions could provide a scalable, data-driven solution to optimise daily light exposure in naturalistic settings.

The LightSPAN study evaluates a chronotherapeutic, mHealth-enabled behaviour change intervention aimed at optimising daily light exposure patterns and supporting circadian health, to facilitate and promote healthy ageing among community-dwelling older adults in Singapore.

## Methods

### Objectives

The general objective of this protocol is to evaluate the effectiveness of the LightSPAN mHealth intervention in optimising light exposure behaviour and improving physiological outcomes among community-dwelling older adults in Singapore.

The specific objectives are:

1. To assess the effectiveness of the LightUP app in optimising daily light exposure patterns
2. To determine whether the intervention improves circadian rest-activity rhythms, sleep quantity and quality, mood and cognitive performance
3. To evaluate effects on frailty status, physical activity, body composition and vitamin D status
4. To examine feasibility, acceptability, usability and user satisfaction of the intervention
5. To explore the acceptability and user experience of the ActLumus light logger device

### Study design

The LightSPAN trial is a randomised, double-blind, parallel-group, placebo-controlled trial conducted among community-dwelling older adults in Singapore. The trial is conducted at Lions Befrienders Active Ageing Centres across Singapore, which serve as community hubs for older adults.

Following eligibility screening and informed consent, participants undergo a 4-week baseline observation period during which habitual light exposure, sleep, and activity patterns are monitored using wearable devices. Participants are then randomised in a 1:1 ratio to either the intervention arm, which receives the LightUP app with active behaviour change features, or the control arm, which receives a placebo version of the app without behavioural components. The intervention period lasts 12 weeks and is followed by a 12-week post-intervention follow-up period, including a 2-week monitoring phase. Study staff conducting outcome assessments are blinded to group allocation throughout the study.

All assessments and app-based interventions are coordinated from TUMCREATE Ltd. Participants are expected to attend assessments at the respective Active Ageing Centres, while the intervention itself is delivered in participants’ home environments using smartphones and wearable devices. Participating Active Ageing Centres provide accessible facilities for participant recruitment and assessment. Personnel involved in delivering the intervention and conducting assessments are trained research staff with specific expertise in cognitive, mood, frailty, and physical performance testing, ensuring consistency and quality in data collection.

The trial design therefore allows for evaluation of both short-term (during intervention) and sustained (post-intervention) effects on light exposure, circadian rhythms, sleep, mood, cognition, frailty, physical activity and vitamin D status. This protocol adheres to the Standard Protocol Items: Recommendations for Interventional Trials (SPIRIT) 2025 guidelines to ensure complete and transparent reporting^18^.

### Patient and public involvement

Members of the public were involved in the design and planning of this study. Feedback from older adults recruited through community partners in Singapore helped to shape the study aims, ensure the intervention content was accessible and adapt the study procedures for cultural and linguistic appropriateness. Input was obtained on the usability of the mobile health app, the acceptability of wearing devices (light logger and activity tracker) and the burden of questionnaires. Community organisations and social service agencies such as Lions Befrienders also contributed to the development of recruitment strategies at Active Ageing Centres and will assist in the dissemination of study findings to participants and the wider community. The process of obtaining feedback from older adults and community partners has been reported in the published co-design protocol^19^. Participants and public representatives were not involved in drafting the present protocol.

### Participants

Participants are community-dwelling older adults aged 60 years and above, residing in Singapore. At the screening visit, participants provide sociodemographic and background information using structured questionnaires. This includes age, sex, highest level of education completed, occupational history, marital status, living arrangement, and smartphone ownership and use, including the smartphone model used during the study. Health history and comorbidity status are assessed using the Self-administered Comorbidity Questionnaire (SCQ)^20^. These screening data are collected to describe the study sample and to support secondary and exploratory analyses examining factors that may influence adherence and intervention outcomes. Eligibility criteria are outlined below.

### Eligibility criteria

Participants must be capable of independent mobility, with or without assistive devices and demonstrate functional independence as assessed by the Lawton Instrumental Activities of Daily Living (IADL) scale^21^, with a score greater than 8 for females and greater than 5 for males. Participants are required to own a smartphone, be proficient in English reading and communication and not be currently enrolled in another mobile health interventional study.

Exclusion criteria include the presence of cognitive impairment, defined as a Montreal Cognitive Assessment (MoCA)^22^ score below 26 after education adjustment, or depressive symptoms, defined as a score greater than 6 on the short form of the Geriatric Depression Scale (GDS)^23^. Individuals with significant physical impairments and/or medical diagnoses that interfere with daily activities, severe terminal illness, or psychiatric conditions affecting daily function are also excluded. Participants with visual impairment, diagnosed eye diseases, ocular abnormalities (as assessed by a licensed optometrist), or uncorrected hearing impairment are not eligible.

### Recruitment, randomization, blinding and treatment allocation

#### Recruitment

Participants will be recruited from Lions Befrienders Active Ageing Centres across Singapore, which provide access to a diverse population of community-dwelling older adults. Recruitment strategies include on-site information sessions, flyers and outreach via community staff. Interested individuals will be screened for eligibility by trained research staff.

Enrolment will continue until the target sample size of 90 participants is reached. Research staff will maintain regular contact with participants during the baseline monitoring period to encourage retention and to address technical issues promptly. The collaboration with established community organisations is expected to facilitate efficient recruitment and reduce participant burden.

#### Randomisation

##### Sequence generation

Randomisation will be conducted using computer-generated block randomisation with the R package *blockTools*, stratified by age, sex and recruitment location to ensure balanced allocation across key demographic variables.

##### Allocation concealment and implementation

To guarantee allocation concealment, the allocation sequence will be securely implemented by a research team member who is not in direct contact with the participants.

Participant enrolment is conducted by trained research staff at the partner Active Ageing Centres. Group assignment is performed only after eligibility confirmation and completion of baseline observation phase. The random allocation sequence is generated by an independent statistician and stored within a secure, central, web-based randomisation module managed by the data management team. Enrolling staff request assignment through this system; the system releases the allocation only at the point of randomisation. Personnel responsible for enrolment and those delivering study procedures have no access to the randomisation list or block details. The master sequence file is access-restricted (statistician and data manager only) and kept on an encrypted server with audit logs.

#### Blinding

##### Who is blinded

This study follows a double-blind outcome-assessor design with participant blinding procedures in place. Both participants and researchers will be blinded to the group assignments to prevent bias. Any technical or logistical queries regarding the use of light loggers and the LightUP app will be addressed by a designated technical support team that is also blinded to the group assignments. This arrangement will reduce the risk of unintentional unblinding through routine interactions and ensure that study integrity is preserved.

##### How blinding is achieved and maintained

Blinding is maintained by using two apps with identical appearance, naming, iconography, onboarding flow and data entry screens. Both apps collect the same light, sleep and mood data and interact identically with the ActLumus logger and Garmin tracker; only the intervention app contains the active behaviour-change features (feedback, goals, prompts), which are implemented via server-side logic not visible to participants or assessors. App store/package identifiers and version numbers are masked in participant-facing materials; installation is performed via a study link that deploys the correct configuration automatically. Staff who perform outcome assessments have no role in app provisioning. To evaluate blinding integrity, participants and assessors will be asked at the endpoint to guess the assigned group and rate their confidence.

##### Unblinding procedures

Unblinding will only be conducted in exceptional cases where it is necessary for safety or technical reasons:

- Medical emergencies: In the event of a medical emergency where knowledge of the group assignment is essential for participant safety, the principal investigator (PI) will be notified. If deemed necessary, the PI will authorize a designated unmasked study personnel to access the randomization code by referring to the secure manual randomization list.
- Technical issues: Should significant technical issues arise with the LightUP app or light loggers, unblinding may be required to resolve the problem. Only the technical team, authorized by the PI, will be granted access to the randomization code to address the issue. Participants will remain blinded throughout this process and no data will be compromised as a result of technical unblinding.

### Informed consent

Written informed consent will be obtained from all participants prior to any study procedures. Trained research staff at the Active Ageing Centres will explain the purpose, procedures, risks and potential benefits of the study using a standardised participant information sheet. Consent will be obtained in English, Mandarin or local dialects, depending on participant preference, and adequate time will be provided for participants to ask questions. Only participants who provide written, signed informed consent will be enrolled in the study.

Consent will cover the collection and use of self-report data, wearable device–derived light and activity data, cognitive performance measures and dried blood spot samples. Participants will also be asked for permission for their anonymised data to be used in future ancillary studies and for sharing under open-access conditions (CC-BY). No biological samples will be stored beyond the planned vitamin D analyses.

### Intervention

#### Experimental intervention

The experimental intervention is the LightUP mHealth smartphone application, which is downloaded onto participants’ own smartphones and used for 12 weeks, following a 4-week baseline monitoring phase. Participants in the intervention group also wear a pendant-worn ActLumus light logger and a wrist-worn Garmin Vivosmart 5 activity tracker throughout the intervention period.

The LightUP app delivers evidence-based behaviour change techniques, including personalised feedback, self-monitoring, goal setting, and educational messages related to healthy light exposure. Participants are able to view their daily light exposure levels directly within the app, supporting self-monitoring and awareness of light-related behaviours. The app was co-designed with older adults and community service providers using a participatory, user-centred approach. The co-design process and development framework have been described in a previously published protocol paper^19^.

Data from the ActLumus light logger are integrated into the app to tailor recommendations and to support participants in meeting daily and weekly light exposure targets. The app incorporates simple reward features, such as the awarding of trophies, to reinforce engagement and goal attainment. To support sustained engagement, participants also receive a mid-intervention drop-in session during the intervention period.

#### Control intervention

The comparator is a placebo version of the LightUP app, which collects light, sleep, and mood data but does not provide behaviour change components such as personalised feedback, prompts, or goal-setting features. Instead, the placebo version retains the same data logging functions and presents neutral content and generic notifications to maintain engagement without directing behaviour.

Both the intervention and placebo versions share a similar interface, navigation structure, and notification timing, ensuring a comparable user experience across groups. Participants in the comparator group also wear the ActLumus light logger and Garmin Vivosmart 5 activity tracker, ensuring equivalent monitoring between groups and isolating the behavioural components of the intervention. Following the 12-week intervention period, participants in both groups enter a 12-week follow-up phase, which includes 2 weeks of extended monitoring.

##### Rationale for comparator

We selected a placebo version of the LightUP app as the comparator to ensure blinding and to isolate the specific effects of the behavioural intervention. Both groups use the same wearable devices (light logger and activity tracker) to monitor light, activity and sleep, but only the intervention group receives active features including personalised feedback, prompts and education. This design allows us to control potential confounding effects of self-monitoring and study participation while testing whether the behavioural features of the LightUP app confer additional benefit. As no standard of care currently exists for promoting light exposure in community-dwelling older adults, a placebo app was chosen over usual care to enhance methodological rigour.

#### Criteria for discontinuing or modifying allocated interventions

Participants may withdraw from the allocated intervention at any time, either at their own request or if adverse events, discomfort, or technical difficulties occur. In such cases, participants are invited to continue follow-up assessments unless they withdraw consent entirely. The intervention protocol is standardised and no modifications to the app are anticipated beyond its automated, data-driven feedback features. All instances of discontinuation or protocol deviation will be recorded.

#### Strategies to improve adherence

Adherence is supported through multiple strategies. The LightUP app incorporates reminders, progress-tracking features and personalised prompts to encourage consistent use. Research staff provide technical support for device setup, troubleshooting and ongoing use, including in-person onboarding with the wearable devices and LightUP installation, in-app tutorial, and a printed step-by-step participant booklet. Ongoing assistance is available via WhatsApp or phone, with in-person support provided on a needs basis. In addition, cloud-based data streams are monitored to identify issues such as failed syncing, battery depletion or limited app interaction, enabling timely follow-up.A mid-intervention drop-in session is conducted to maintain engagement. Overall, adherence and quality control of device and app-related components is monitored through app usage logs, wearable device data and completion of scheduled assessments.

#### Concomitant care

Participants are permitted to continue all aspects of their usual medical care, daily routines and lifestyle activities throughout the trial. However, concurrent participation in other mobile health intervention studies or trials targeting circadian rest-activity rhythms, sleep, mood, cognition, frailty, physical function, or vitamin D is not permitted, to avoid contamination and ensure that behavioural changes can be attributed to the LightUP app. All relevant changes in health status or treatments during the study will be documented.

### Outcomes

#### Primary outcome

The primary outcome is the effectiveness of the LightUP app in improving light exposure behaviour in older adults. This will be assessed using data from the ActLumus wearable light logger (ActLumus, Condor Instruments; São Paolo, Brazil) across the 4-week baseline, the 12-week intervention and the 2-week follow-up monitoring period. The primary analysis metric is the mean daily time (minutes per day) spent above 250 lx melanopic equivalent daylight illuminance (melanopic EDI) during daytime hours. Changes will be calculated relative to baseline and both short-term effects (during the 12-week intervention) and longer-term effects (during follow-up) will be examined.

#### Secondary outcomes

Secondary outcomes span multiple domains.

#### Sleep

Quantity and quality: total sleep time, sleep efficiency, sleep onset latency and wake after sleep onset, measured via Garmin Vivosmart 5 (Garmin Ltd., Olathe, KS, USA) actigraphy and sleep diaries throughout the study.

Subjective quality: assessed using the Pittsburgh Sleep Quality Index (PSQI)^24^ at weeks 4 (baseline), 10 (midpoint), 16 (endpoint) and 28 (follow-up).

#### Circadian rhythms

Interdaily stability, intradaily variability and relative amplitude of rest-activity cycles, measured continuously with Garmin data across the study period.

#### Mood

Mood will be assessed using the Brief Mood Introspection Scale (BMIS)^25^ at baseline, midpoint, endpoint and follow-up. Daily mood will be captured using the Single-Item Mood Scale (SIMS)^26^. Weekly mood ratings will be assessed using the Visual Analogue Mood Scale (VAMS) during the intervention.

#### Cognition

Cognitive performance will be assessed using the NIH Toolbox® battery^27^ at baseline, midpoint, endpoint and follow-up, including the Dimensional Change Card Sort, Flanker Inhibitory Control and Attention, Oral Symbol Digit, Picture Sequence Memory, Rey Auditory Verbal Learning and Pattern Comparison Processing Speed tests.

#### Frailty and physical activity

Frailty will be assessed using standard criteria (weight loss, exhaustion, low activity via Physical Activity Scale for the Elderly (PASE), slowness via 5-metre walk and grip strength^28^). Physical activity outcomes including daily step count, sedentary behaviour, moderate-to-vigorous activity will be assessed using Garmin wearable data.

#### Body composition

Body composition outcomes, including weight, body fat percentage, muscle mass, bone mass and body mass index (BMI), were assessed at baseline, midpoint, endpoint and follow-up using a bioelectrical impedance analysis (BIA) machine (TANITA DC-430MA, Tanita Corporation, Tokyo, Japan).

#### Vitamin D status

Vitamin D status will be assessed from dried blood spot samples at the same timepoints. 25-hydroxyvitamin D [25(OH)D] concentrations will be measured with a dried blood spot test kit (Vitamin D Test, NeoVos by SureScreen Health, Newcastle Upon Tyne, United Kingdom).

#### Feasibility, usability and acceptability

Usability of the LightUP app measured by the mHealth App Usability Questionnaire (MAUQ)^29^ at endpoint. Acceptability of the app and wearable devices assessed using an adapted Technology Acceptance Model (TAM) questionnaire^30^ at endpoint. Qualitative feedback on participant experience, app usage behaviour and overall acceptability of the app will be obtained through a focus group discussion at endpoint assessment.

#### Time points for analysis

Primary and secondary outcomes will be analysed at baseline (week 4), mid-intervention (week 10), end of intervention (week 16) and follow-up (week 28), depending on the measure. Continuous monitoring outcomes (e.g. light exposure, actigraphy, mood diaries) will be aggregated into daily or weekly metrics as appropriate.

### Participant timeline

The participant timeline and flow through the study are visualised in *Supplementary Figure 1.* Following eligibility screening, obtaining informed consent, and documenting sociodemographic and heath characteristics at Week 1, participants undergo a 4-week baseline monitoring period (weeks 1-4). During this period, they wear the ActLumus light logger and Garmin Vivosmart 5 activity tracker.

Following the baseline monitoring period, participants undergo baseline assessments (week 4) and are randomised in a 1:1 ratio to either the intervention or comparator arm.

The intervention phase lasts 12 weeks (weeks 4-16). Participants in the intervention group use the LightUP mobile health app, which delivers personalised feedback, education and goal setting, while participants in the comparator group use the placebo version of the app that records but does not provide feedback. Both groups continue to wear the ActLumus logger and Garmin device throughout this period. An additional drop-in session is delivered at week 10 to reinforce adherence.

A follow-up period of 12 weeks (weeks 17-28) follows the intervention. During this time, participants no longer use the app but continue their usual monitoring and assessments. The follow-up includes a 2-week monitoring phase with both the ActLumus and Garmin devices to assess sustained changes in light exposure, sleep and circadian rhythms.

Assessments are scheduled at four key time points:

- **Week 4 – Baseline assessment (BL):** sleep quality (PSQI), mood (BMIS), cognition (NIH Toolbox), frailty (five criteria), physical activity (PASE), body composition, vitamin D (25(OH)D).
- **Week 10 – Mid-point assessment (MP):** interim assessments of sleep, mood, cognition, frailty, physical activity, body composition and vitamin D.
- **Week 16 – End-point assessment (EP):** repeat assessments including usability (MAUQ), acceptability (TAM) questionnaires and qualitative post-trial discussion.
- **Week 28 – Follow-up assessment (FU):** repeat full assessments, including 2 weeks of device-based monitoring.

### Data collection

Outcome data will be collected using a combination of wearable devices, validated questionnaires, cognitive tests and clinical assessments as outlined in the Outcomes section of this protocol. Light exposure is measured continuously using the ActLumus pendant logger, while rest-activity cycles, sleep and physical activity are recorded using the Garmin Vivosmart 5 tracker. As outlined in Outcomes, validated self-report instruments include the PSQI, BMIS, VAMS, NIH Toolbox cognitive tests and PASE. Frailty measures include grip strength, gait speed, weight loss, exhaustion and activity level. Body composition is assessed using BIA and vitamin D is measured using dried blood spot analysis. Usability and acceptability of the intervention are assessed using the MAUQ and an adapted TAM survey.

To ensure quality, all assessors are trained and certified in administering the questionnaires and performance-based measures. Standard operating procedures are in place for device calibration, bio sample handling and data transfer. Data is collected electronically using secure, tablet-based entry systems, with automatic checks for out-of-range or missing values.

Participant retention and completeness of data collection will be supported through regular check-ins, technical support for device use and reminder notifications via the app in both trial arms. Participants who discontinue the intervention will still be invited to complete outcome assessments at scheduled visits unless they withdraw consent entirely. Reasons for non-adherence (e.g. device discomfort, loss of interest) or non-retention (e.g. withdrawal, loss to follow-up) will be systematically documented.

### Data management

All study data will be entered into a secure, password-protected electronic database hosted on institutional servers. Data entry will be performed electronically, with built-in range checks, mandatory fields and double verification procedures for critical variables. Data from wearable devices and the mobile application will be automatically synchronised with the study server via encrypted data transfer. Bio sample results will be entered by laboratory staff following validation procedures.

Participant confidentiality will be preserved throughout the trial. Personal identifiers (e.g. name, contact details) will be collected separately from research data and stored in encrypted, access-restricted files accessible only to authorised study staff. Each participant will be assigned a unique study identification number, which will be used for all data collection and analysis. Wearable device and app data will be encrypted at rest and in transit and stored in compliance with the Singapore Personal Data Protection Act. All data shared with collaborators or made publicly available will be fully anonymised.

Data security will be ensured through role-based access controls, password-protected user accounts and regular system backups. The final locked dataset will be archived on secure institutional servers for a minimum of 10 years following study completion.

### Sample size

Based on power calculations conducted using existing light exposure data, anchored to the primary outcome of time spent above threshold (TAT), defined as time spent above 250 lx melanopic EDI during daytime (TAT_250_), we estimate that a sample size of 25 participants per group (i.e., 50 participants in total) is sufficient to detect a 20% change in light exposure metrics with 80% power, assuming a standard deviation of 0.2 and a two-sided alpha of 0.05.

These calculations were based on an existing dataset from Malaysia, derived from a previous study comparing light exposure patterns in Switzerland and Malaysia^31^. This dataset is the closest available approximation to our target study population in terms of geographic and lifestyle similarities. The sample size estimation is documented in full in the project GitHub repository (https://github.com/tscnlab/MurukesuEtAl_BMCGeriatr_2026; https://doi.org/10.5281/zenodo.17914996 archived on Zenodo)

In the Malaysian dataset, daily time spent above 250 lx melanopic EDI (TAT_250_) averaged approximately 98 minutes per day, with 490 daily observations nested within 19 participants. This hierarchical data structure was retained in the power analysis using a mixed-effects model specification of the form outcome ∼ intervention + (intervention | participant ID). As the original dataset was modelled using a Poisson distribution, the same distributional assumption was applied when generating simulation data for the power analysis via bootstrapping.

The power analysis focused on three light exposure metrics derived from the Malaysia dataset, as these represent conceptually relevant aspects of daytime and evening light exposure targeted by the LightSPAN intervention:

- Time spent above 1000 lx melanopic EDI during the daytime (TAT_1000_)
- Time spent above 250 lx melanopic EDI during the daytime (TAT_250_)
- Time spent below 10 lx melanopic EDI in the evening (TBT_10_)

All three metrics are relevant in the context of the project, with TAT_250_ designated as the primary outcome. Importantly, the required sample sizes were identical for all three metrics when evaluated in increments of five participants per group, and therefore the same sample size estimate applies to the primary outcome.

A 20% relative change, which defines the minimum detectable effect for the primary outcome (TAT_250_), in each of these metrics corresponds to approximately:

- ∼9 minutes/day increase above 1000 lx melanopic EDI
- ∼20 minutes/day increase above 250 lx melanopic EDI
- ∼30 minutes/day reduction below 10 lx melanopic EDI in the evening

However, to account for:

- Additional outcome domains such as sleep, mood and cognitive function, which may show greater variability or require larger sample sizes to detect meaningful effects
- Considering the intensive nature of the trial and to effectively account for potential dropouts, an expected attrition rate of 25-30%, which is typical in studies involving older adults
- The need for more robust, generalizable results across a wider population

We plan to recruit a total of 90 participants (45 per group). This allows for approximately 63-68 participants to complete the study, corresponding to 31-34 participants per group, even after accounting for attrition. The recruitment strategy is guided by findings from similar mHealth interventions involving older adults, where dropout rates tend to be higher due to factors such as technology challenges, health status and varying levels of digital literacy. Studies in this demographic^32^ ^33^ have demonstrated that over-recruiting by approximately 25-30% is a best practice to ensure adequate sample sizes for meaningful data analysis. By aiming to retain at least 90 participants, we align with research standards that suggest this number is sufficient for exploratory analysis of the intervention’s effectiveness and feasibility.

### Statistical analysis

#### Analysis population

Analyses will follow the intention-to-treat principle, including all randomised participants in their assigned groups. A per-protocol sensitivity analysis will be performed including only participants with high adherence (≥80% app engagement and valid device wear).

#### Primary outcome analysis

The primary endpoint is light exposure behaviour, operationalised as the mean daily time (minutes per day) spent above 250 lx melanopic EDI during daytime hours. Light exposure patterns will be analysed per individual using generalized linear mixed models (GLMMs). Daily TAT values will be entered into models including group assignment and assessment block/day-in-trial as fixed effects, with participant-level random effects to account for repeated daily measurements.

Based on prior analyses of TAT in a Malaysia–Switzerland light exposure study^31^, a Poisson error distribution was identified as an appropriate fit for mixed-effects modelling and is therefore used for all analyses with TAT as the dependent variable.

The full model in Wilkinson notation will follow this specification:

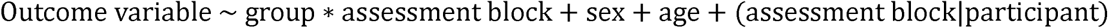

Model diagnostics will include visual checks for homoscedasticity, normality of residuals and random effects, and their location at zero. Baseline characteristics, including sociodemographic variables, health status, and baseline outcome measures, will be summarised descriptively by study group, and selected variables may be included as covariates in secondary or exploratory analyses where appropriate.

#### Secondary outcomes analyses

Secondary endpoints, as defined in SPIRIT item 16, include outcomes spanning sleep, circadian rhythms, mood, cognition, frailty, physical activity, body composition, vitamin D status, and feasibility-related measures, assessed at BL, MP, EP, and FU, depending on the measure.

Quantitative differences across assessment time points will be examined using GLMMs or linear mixed-effects models, depending on the distributional properties of each outcome. Models will include group, assessment block, and group-by-assessment block interaction as fixed effects, with participant included as a random effect to account for repeated measures.

Statistical significance will be assessed using a two-sided alpha level of 0.05. p-values will be calculated using likelihood ratio tests comparing models with and without the parameter of interest. Unstandardised effect sizes (beta coefficients) will be reported. A false discovery rate correction will be applied within each branch of secondary outcome analyses, based on the number of models or tests performed within that branch.

#### Handling of missing data

Missing outcome data will be examined for patterns and assumed to be missing at random. Mixed-effects models inherently accommodate unbalanced repeated measures. For questionnaire outcomes, multiple imputations using chained equations will be considered. Sensitivity analyses (best-case/worst-case) will be conducted to assess robustness of findings. Missing or irregular data from the wearable light loggers will not be imputed. Irregular data may be regularized to a consistent time series of data within a participant and collection period. Participant days missing more than 2 hours of data will be excluded from metric calculations and thus further analysis. Biller et al. (Malaysia/Switzerland) found that 6 hours of missing data led to non-significant differences in a month-long data collection effort per participant^31^. As the follow-up period only contains 2 weeks of data collection, we choose this more conservative threshold.

#### Additional and exploratory analyses

Exploratory analyses will incorporate baseline, midpoint, endpoint, and follow-up as covariates or additional outcomes to characterise temporal patterns beyond the primary and secondary endpoints. Bayesian analyses may be conducted as a complementary approach, providing posterior estimates with corresponding credible intervals.

Continuously collected measurements, such as light exposure, might further be explored non-linearly through generalized additive mixed models (GAMMs)^34^, which have been shown to be a good fit for personal light exposure data^35^. Additional exploratory analyses will be pre-registered on the Open Science Framework prior to execution to ensure transparency.

#### Interim analyses and stopping guidelines

No formal interim analyses are planned. Stopping criteria will be limited to safety concerns identified by the Steering Committee (e.g., unforeseen adverse events related to device use). Any decision to terminate or modify the trial will rest with the Principal Investigator in consultation with the Steering Committee and the Institutional Review Board (IRB).

#### Statistical software

Statistical analyses will be conducted using R. Particular R packages that are planned to be used, among others, include *LightLogR*^36^ for processing and calculating light data and variables, and *tidyverse*^37^ for a stringent, tidy analysis framework.

### Adverse events

The LightSPAN trial is considered minimal risk, as the intervention involves the use of a mHealth application and wearable monitoring devices. There are no known serious harms associated with these technologies. Possible risks include minor discomfort from wearing the ActLumus light logger or Garmin Vivosmart 5 activity tracker. These risks are considered low and are comparable to wearing commercially available wearable devices. No invasive procedures are involved except for dried blood spot collection for vitamin D analysis, which may cause minor discomfort.

#### Systematically assessed adverse events

Participants excluded following ocular health screening will receive a referral letter outlining the findings and recommending follow-up with an appropriate healthcare provider or specialist.

#### Non-systematically assessed adverse events

Participants are encouraged to report any problems or adverse experiences spontaneously to the study team throughout the trial. Reports will be collected via phone, email, or in person during centre visits.

#### Coding and grading of adverse events

All adverse events will be documented and coded by study staff according to severity (mild, moderate, severe) and relatedness to the intervention (unrelated, possibly related, probably related). Coding will be performed by personnel blinded to group allocation. No formal clinical grading system is planned, as no serious medical harms are anticipated.

#### Grouping of harms and reporting of adverse events

Harms will be grouped by seriousness (serious versus non-serious), severity and type (e.g., device-related discomfort, app-related difficulties). Any adverse events leading to discontinuation of the intervention will be recorded separately. If a serious adverse event is reported, it will be immediately reviewed by the Principal Investigator and reported to the Parkway Independent Ethics Committee (PIEC), in line with ethical approval requirements.

### Ancillary and post-trial care

As the intervention involves a digital health app and wearable devices, no specific ancillary or post-trial medical care is anticipated. Any discomfort related to device wear (e.g. skin irritation) will be addressed by study staff. Participants will be covered by institutional insurance policies for study-related harm, in accordance with local regulations. No compensation is planned for non-study-related health events. Participants will receive a lay summary of the study findings at trial completion.

### Audits and inspections

#### Composition and role

Given the minimal-risk nature of the intervention (digital health app and wearable devices), no independent Data Monitoring Committee (DMC) will be established. Trial oversight will be managed by the study’s Steering Committee, which includes investigators from TUMCREATE and collaborating institutions. The Steering Committee meets monthly to review recruitment, adherence, data quality and any adverse events.

#### Trial monitoring

Trial conduct will be monitored by the institutional research governance office, which will conduct regular remote monitoring of recruitment, consent forms and data quality. Monitoring will include verification of consent, adherence to protocol and reporting of adverse events. No on-site audits are anticipated unless concerns arise.

### Protocol amendments

All substantial protocol amendments (e.g., changes to eligibility criteria, outcomes, analyses) will be reviewed and approved by the relevant IRBs prior to implementation. Updates will also be made in the trial registry (ISRCTN12391932, registered 5 September 2025) and communicated to study sites, funders and participants as appropriate. Minor administrative amendments (e.g., corrections of typographical errors) will be documented in the trial master file.

## Discussion

### Strengths

This study has several strengths. It represents a novel digital behavioural intervention that specifically targets environmental light exposure as a modifiable, lifestyle-based strategy for preventive health and health promotion among older adults. To our knowledge, this is the first study to operationalise light exposure optimisation through a mobile health platform in this population. The trial is underpinned by a rigorous methodological design, employing a double-blind randomised controlled framework with allocation concealment and stratified balancing by sex, age, and recruitment site. These design features minimise selection and performance bias and enhance internal validity. A further strength is the use of a multi-modal assessment framework that combines objective measures of light exposure, physical activity, and physiological outcomes, captured using the ActLumus light logger and Garmin Vivosmart 5, with validated subjective instruments. This approach enables comprehensive outcome assessment and robust triangulation across measurement modalities. The inclusion of a pilot and feasibility phase allows refinement of study procedures, identification of implementation challenges, and optimisation of intervention delivery prior to the main trial, thereby reducing operational risks and strengthening overall feasibility. Finally, recruitment through Lions Befrienders’ Active Ageing Centres situates the intervention within a real-world community context, enhancing ecological validity and supporting the potential translation of findings into community-based and public health programmes.

### Limitations

Several limitations should be considered. The digital nature of the intervention may impose a technology-related burden for some older participants, particularly with respect to device wear, data synchronisation, and mobile application use, which could affect adherence and data completeness. Furthermore, sample size assumptions were informed by effect size estimates derived from a Malaysian dataset, which may not fully reflect variability within the Singaporean older adult population and may introduce uncertainty in power estimation. Although over-recruitment has been planned, attrition related to digital literacy, participant burden, or device fatigue remains a potential risk and could reduce effective statistical power. In addition, the exclusion of individuals with cognitive impairment, ocular disease, or clinically significant depressive symptoms was necessary to ensure participant safety and reduce clinical and behavioural confounding. However, this may limit generalisability, resulting in a study sample that is comparatively healthier and more digitally capable than the broader older adult population. While blinding is strengthened by the identical visual design and navigation of the intervention and placebo applications, functional differences between versions may allow some participants to infer group allocation. Although outcome assessors remain blinded, the possibility of partial participant unblinding and residual performance bias cannot be entirely excluded.

### Publications and dissemination policy

The findings of this study will be primarily disseminated through scientific channels. Results will be published in peer-reviewed open access journals and preprints will be deposited on servers such as medRxiv to allow rapid access by the research community. In addition, anonymised participant-level data, statistical code and supporting documentation will be deposited in an open repository to ensure transparency and facilitate secondary analyses.

Authorship will follow International Committee of Medical Journal Editors (ICMJE) guidelines and no professional medical writers will be involved in the preparation of manuscripts.

#### Protocol and statistical analysis plan

The full trial protocol will be published in *BMC Geriatrics*. Any protocol amendments will be updated in the ISRCTN record to ensure accuracy and transparency. A separate statistical analysis plan will be finalised before database lock and will be made openly available on the Open Science Framework (OSF). This approach ensures that analytical procedures are defined in advance and can be independently scrutinised.

### Trial status

Recruitment for the LightSPAN trial commenced on 18 August 2025. The trial is currently ongoing and is expected to continue until 30 November 2026. The trial is registered with the International Standard Randomised Controlled Trial Number registry under identifier ISRCTN12391932.

## List of abbreviations

BIA: Bioelectrical impedance analysis
BL: Baseline
BMI: Body mass index
BMIS: Brief Mood Introspection Scale
CC-BY: Creative Commons Attribution license
CIE: International Commission on Illumination
DBS: Dried blood spot
DMC: Data Monitoring Committee
EDI: Equivalent daylight illuminance
EP: Endpoint
FU: Follow-up
GAMM: Generalised additive mixed model
GDS: Geriatric Depression Scale
GLMM: Generalised linear mixed model
IADL: Instrumental Activities of Daily Living
IRB: Institutional Review Board
ISRCTN: International Standard Randomised Controlled Trial Number
MAUQ: mHealth App Usability Questionnaire
mHealth: Mobile Health
MoCA: Montreal Cognitive Assessment
MP: Midpoint
NIH: National Institutes of Health
OSF: Open Science Framework
PASE: Physical Activity Scale for the Elderly
PI: Principal Investigator
PIEC: Parkway Independent Ethics Committee
PSQI: Pittsburgh Sleep Quality Index
RCT: Randomised controlled trial
SCQ: Self-administered Comorbidity Questionnaire
SIMS: Single-Item Mood Scale
SPIRIT: Standard Protocol Items: Recommendations for Interventional Trials
TAM: Technology Acceptance Model
TAT: Time above threshold
TAT250: Time spent above 250 lx melanopic equivalent daylight illuminance
TBT10: Time spent below 10 lx melanopic equivalent daylight illuminance
VAMS: Visual Analogue Mood Scale
25(OH)D: 25-hydroxyvitamin D

## Declarations

### Ethics approval and consent to participate

The study protocol, informed consent form and study materials have received approval from the Parkway Independent Ethics Committee (ref PIEC/2024/041, approved 18 June 2025). The protocol is also registered at the ISRCTN – The UK’s Clinical Study Registry on 5 September 2025 (ID: ISRCTN123919320, https://doi.org/10.1186/ISRCTN12391932). Written informed consent will be obtained from all participants prior to any study procedures. Any modifications will be submitted for ethics review prior to implementation.

### Consent for publication

Not applicable

### Availability of data and materials

Upon completion of the trial and publication of the primary outcomes, anonymised participant-level data, the accompanying data dictionary, statistical code and all supporting documentation will be made available through the Open Science Framework. Data will be released under a CC-BY license, allowing unrestricted reuse with appropriate attribution. Shared materials will include de-identified data on light exposure, sleep, mood, cognition and vitamin D status, along with the relevant analytical scripts written in R and Python and documentation associated with the intervention. No application process will be required to access the dataset.

### Competing interests

J.Z. declares the following potential conflicts of interest in the past five years (2021-2025). Academic roles: Member of Joint Technical Committee 20 (JTC20) of the International Commission on Illumination (CIE); Member of Research Data Alliance Working Group Optical Radiation and Visual Experience Data; Speaker of group 2 (melanopic effects of light) of the Technical Scientific Committee (TWA) of the German Society of Lighting Technology and Design (LiTG) Remunerated roles: Examiner, Swiss Lighting Society; Teacher, LiTG; Teacher, University of Applied Sciences, Munich, Teacher, Technical University of Applied Sciences, Rosenheim. Associated partner, 3lpi lighting design + engineering, Munich. Tool- and 3D-model design, Zumtobel Lighting GmbH; Course design, University of Applied Sciences, Munich & Virtual University Bavaria. Honoraria for talks: Received honoraria from LiTG; Lamilux (Heinrich Strunz GmbH); Robert-Bosch Hospital Stuttgart; Ergotopia GmbH; German statutory accident insurance institution for the administrative sector (VBG); BRIXEN CULTUR, Italy; KITEO GmbH & Co.KG; University of Applied Sciences Augsburg. Travel reimbursements: Daimler und Benz Stiftung. Patents: Together with 3lpi holds a design patent for non-visually optimized luminaire (No 008194021-0001 through -0006) at the European Union Intellectual property office.

M.S. declares the following potential conflicts of interest in the past five years (2021–2025). Academic roles: Member of the Board of Directors, Society of Light, Rhythms and Circadian Health (SLRCH); Chair of Joint Technical Committee 20 (JTC20) of the International Commission on Illumination (CIE); Member of the Daylight Academy; Chair of Research Data Alliance Working Group Optical Radiation and Visual Experience Data. Remunerated roles: Speaker of the Steering Committee of the Daylight Academy; Ad-hoc reviewer for the Health and Digital Executive Agency of the European Commission; Ad-hoc reviewer for the Swedish Research Council; Associate Editor for LEUKOS, journal of the Illuminating Engineering Society; Examiner, University of Manchester; Examiner, Flinders University; Examiner, University of Southern Norway. Funding: Received research funding and support from the Max Planck Society, Max Planck Foundation, Max Planck Innovation, Technical University of Munich, Wellcome Trust, National Research Foundation Singapore, European Partnership on Metrology, VELUX Foundation, Bayerisch-Tschechische Hochschulagentur (BTHA), BayFrance (Bayerisch-Französisches Hochschulzentrum), BayFOR (Bayerische Forschungsallianz) and Reality Labs Research. Honoraria for talks: Received honoraria from the ISGlobal, Research Foundation of the City University of New York and the Stadt Ebersberg, Museum Wald und Umwelt. Travel reimbursements: Daimler und Benz Stiftung. Patents: Named on European Patent Application EP23159999.4A (“System and method for corneal-plane physiologically-relevant light logging with an application to personalized light interventions related to health and well-being”). The funders had no role in study design, data collection and analysis, decision to publish or preparation of the manuscript. Authors R.R.M., Z.A.A. and D.d.V. do not declare any competing interests.

### Funding

This research is funded by the National Research Foundation, Prime Minister’s Office, Singapore, under its Campus for Research Excellence and Technological Enterprise (CREATE) programme through the project *Optimising environmental light exposure at scale to support eye and brain development and health across the lifespan (LightSPAN)* (NRF2022-THE004-0002). JZ’s position was supported by MeLiDos (project 22NRM05), which has received funding from the European Partnership on Metrology, co-financed by the European Union’s Horizon Europe Research and Innovation Programme and by the Participating States.

TUM CREATE Ltd. is the sponsor of this trial and is responsible for ensuring that the study is conducted in accordance with applicable regulatory and ethical requirements. The sponsor and funders had no role in the study design; data collection, analysis or interpretation; the decision to publish; or the preparation of the manuscript. The views expressed are those of the authors and do not necessarily reflect those of the European Union, EURAMET or the funding bodies.

### Authors’ contributions

MS conceived the study and is the principal investigator, leading the overall trial. RRM, ZAA and DDV contributed to study conception, methods development and protocol drafting. DDV additionally led the development of the research software. JZ contributed to the formal analysis. MS supervised the study and was responsible for funding acquisition. RRM and MS managed project administration. All authors contributed to revising the manuscript, read and approved the final version.

## Supporting information

SPIRIT Checklist

## Acknowledgements

We acknowledge the support of the Lions Befrienders Singapore Active Ageing Centres and the community partners involved in the planning and implementation of this study.

## Structured summary

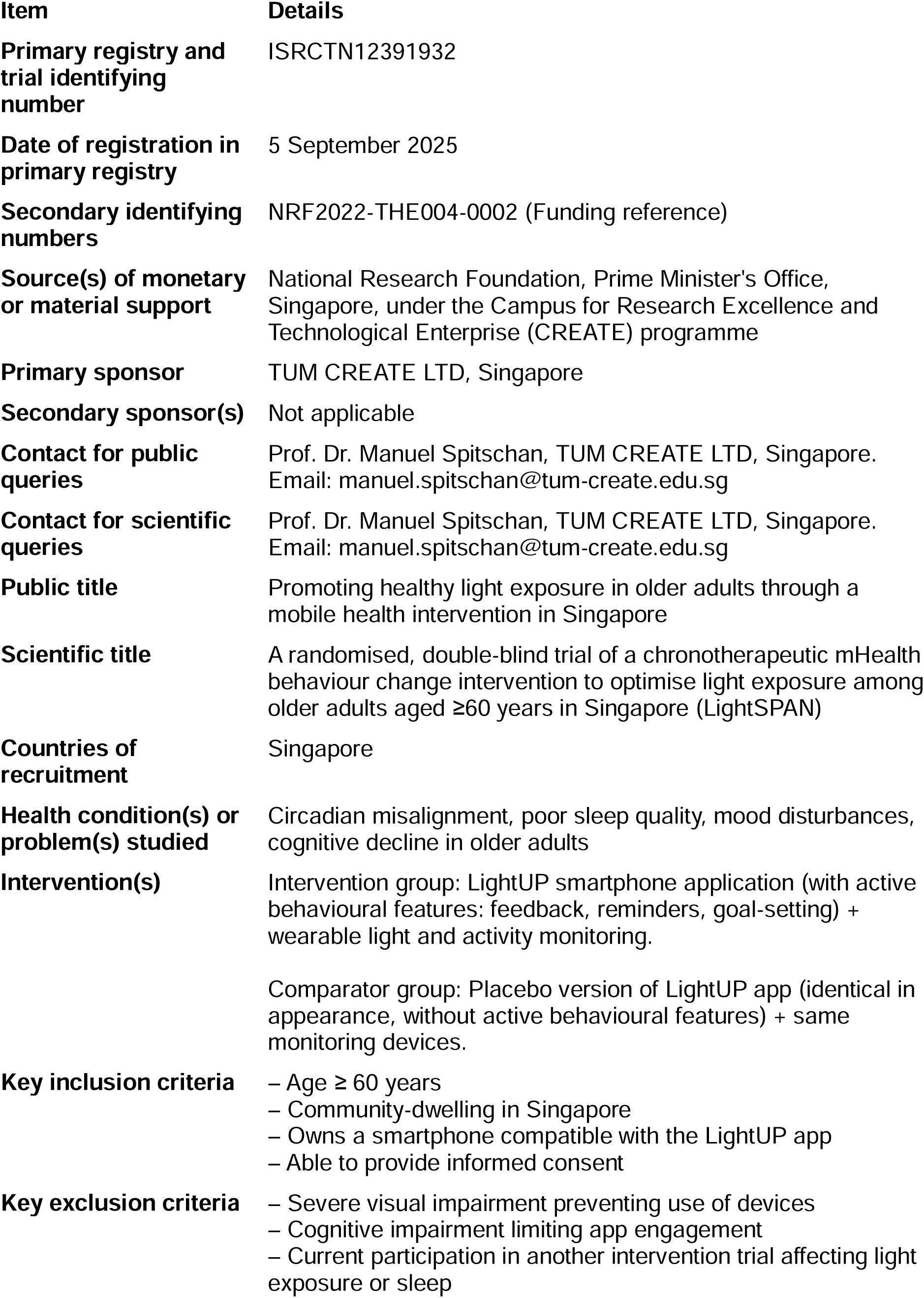
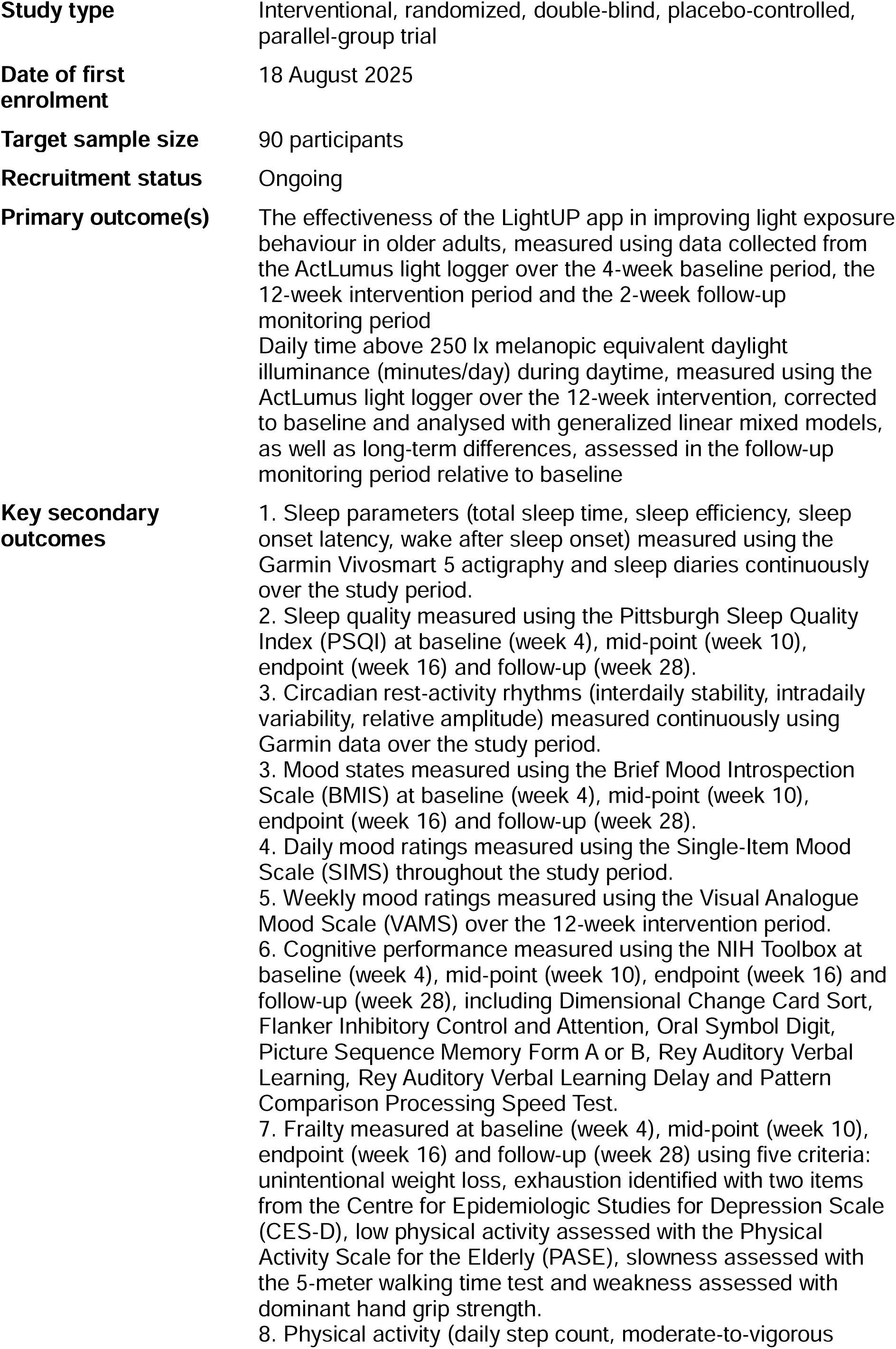
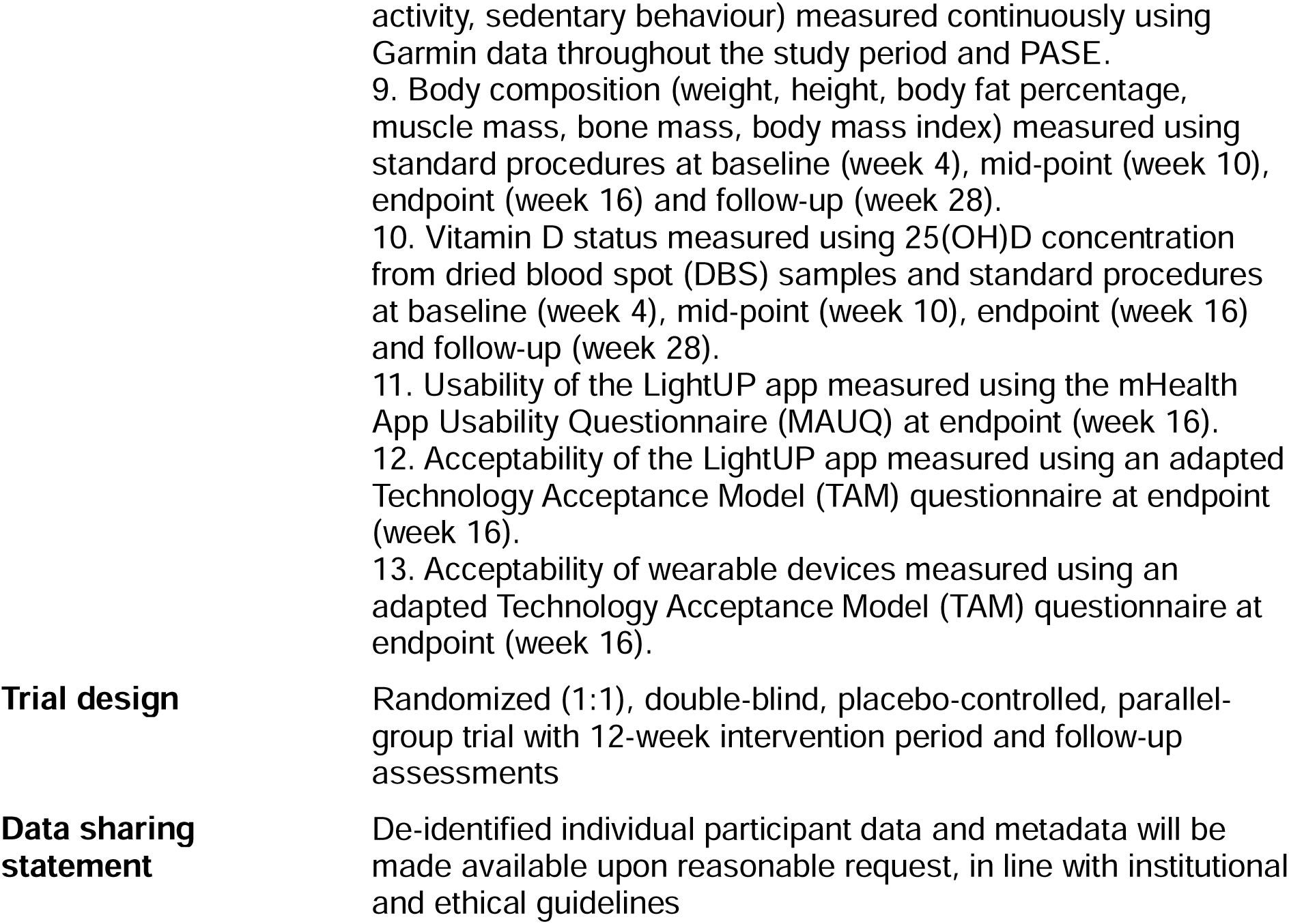

## Notes

### Clinical Trial

ISRCTN12391932

### Funding Statement

This research is funded by the National Research Foundation Singapore through the programme Optimising environmental light exposure at scale to support eye and brain development and health across the lifespan (LightSPAN), reference NRF2022-THE004-0002 and the Max Planck Society.

### Author Declarations

Ethics approval obtained from Parkway Independent Ethics Committee with reference number PIEC/2024/041

### Summary of Updates

In response to the editor and reviewer comments, we revised the manuscript formatting and section placement to align with the BMC Geriatrics protocol format; these changes involved reorganisation and clarification only, with no substantive modifications to the scientific content, study design, or planned analyses. Specifically, we added a Discussion section to both the abstract and the main manuscript as required for protocol papers, relocated the strengths and limitations from bullet points in the abstract to the Discussion, added an Acknowledgements section and a List of Abbreviations, moved the Structured summary to the Supplementary Materials, corrected the recruitment completion date to reflect the updated ISRCTN registry information, and updated the SPIRIT checklist to match the revised structure and information placement.

## References

1. Blume C, Garbazza C, Spitschan M. Effects of light on human circadian rhythms, sleep and mood. Somnologie (Berl) 2019;23(3):147–56. doi: 10.1007/s11818-019-00215-x [published Online First: 20190820]

2. Burns AC, Windred DP, Rutter MK, et al. Day and night light exposure are associated with psychiatric disorders: an objective light study in >85,000 people. Nature Mental Health 2023;1(11):853–62. doi: 10.1038/s44220-023-00135-8

3. Windred DP, Burns AC, Lane JM, et al. Brighter nights and darker days predict higher mortality risk: A prospective analysis of personal light exposure in >88,000 individuals. Proc Natl Acad Sci U S A 2024;121(43):e2405924121. doi: 10.1073/pnas.2405924121 [published Online First: 20241015]

4. Windred DP, Burns AC, Rutter MK, et al. Personal light exposure patterns and incidence of type 2 diabetes: analysis of 13 million hours of light sensor data and 670,000 person-years of prospective observation. Lancet Reg Health Eur 2024;42:100943. doi: 10.1016/j.lanepe.2024.100943 [published Online First: 20240605]

5. Lazar R, Degen J, Fiechter AS, et al. Regulation of pupil size in natural vision across the human lifespan. R Soc Open Sci 2024;11(6):191613. doi: 10.1098/rsos.191613 [published Online First: 20240619]

6. Verma AK, Singh S, Rizvi SI. Aging, circadian disruption and neurodegeneration: Interesting interplay. Exp Gerontol 2023;172:112076. doi: 10.1016/j.exger.2022.112076 [published Online First: 20221224]

7. Dev MK, Black AA, Cuda D, et al. Low Light Exposure and Physical Activity in Older Adults With and Without Age-Related Macular Degeneration. Transl Vis Sci Technol 2022;11(3):21. doi: 10.1167/tvst.11.3.21

8. Hood S, Amir S. The aging clock: circadian rhythms and later life. J Clin Invest 2017;127(2):437–46. doi: 10.1172/JCI90328 [published Online First: 20170201]

9. Lin JB, Tsubota K, Apte RS. A glimpse at the aging eye. NPJ Aging Mech Dis 2016;2:16003. doi: 10.1038/npjamd.2016.3 [published Online First: 20160310]

10. Duffy JF, Zitting KM, Chinoy ED. Aging and Circadian Rhythms. Sleep Med Clin 2015;10(4):423–34. doi: 10.1016/j.jsmc.2015.08.002 [published Online First: 20150915]

11. Owsley C. Aging and vision. Vision Res 2011;51(13):1610–22. doi: 10.1016/j.visres.2010.10.020 [published Online First: 20101023]

12. Van Someren EJ, Kessler A, Mirmiran M, et al. Indirect bright light improves circadian rest-activity rhythm disturbances in demented patients. Biol Psychiatry 1997;41(9):955–63. doi: 10.1016/S0006-3223(97)89928-3

13. Skjerve A, Bjorvatn B, Holsten F. Light therapy for behavioural and psychological symptoms of dementia. Int J Geriatr Psychiatry 2004;19(6):516–22. doi: 10.1002/gps.1087

14. Forbes D, Blake CM, Thiessen EJ, et al. Light therapy for improving cognition, activities of daily living, sleep, challenging behaviour, and psychiatric disturbances in dementia. Cochrane Database Syst Rev 2014;2014(2):CD003946. doi: 10.1002/14651858.CD003946.pub4 [published Online First: 20140226]

15. Faulkner SM, Dijk DJ, Drake RJ, et al. Adherence and acceptability of light therapies to improve sleep in intrinsic circadian rhythm sleep disorders and neuropsychiatric illness: a systematic review. Sleep Health 2020;6(5):690–701. doi: 10.1016/j.sleh.2020.01.014 [published Online First: 20200312]

16. Biller AM, Balakrishnan P, Spitschan M. Behavioural determinants of physiologically-relevant light exposure. Commun Psychol 2024;2(1):114. doi: 10.1038/s44271-024-00159-5 [published Online First: 20241129]

17. Siraji MA, Lazar RR, van Duijnhoven J, et al. An inventory of human light exposure behaviour. Sci Rep 2023;13(1):22151. doi: 10.1038/s41598-023-48241-y [published Online First: 20231213]

18. Chan AW, Boutron I, Hopewell S, et al. SPIRIT 2025 Statement: Updated Guideline for Protocols of Randomized Trials. JAMA 2025;334(5):435–43. doi: 10.1001/jama.2025.4486

19. Alkaff ZA, Murukesu RR, Del Villar D, et al. Protocol for a co-design study for the development of a chronotherapeutic mobile health behaviour change intervention targeting light exposure among older adults. F1000Res 2024;13:1356. doi: 10.12688/f1000research.157814.2 [published Online First: 20250725]

20. Sangha O, Stucki G, Liang MH, et al. The Self-Administered Comorbidity Questionnaire: a new method to assess comorbidity for clinical and health services research. Arthritis Rheum 2003;49(2):156–63. doi: 10.1002/art.10993

21. Lawton MP, Brody EM. Assessment of older people: self-maintaining and instrumental activities of daily living. Gerontologist 1969;9(3):179–86.

22. Nasreddine ZS, Phillips NA, Bedirian V, et al. The Montreal Cognitive Assessment, MoCA: a brief screening tool for mild cognitive impairment. J Am Geriatr Soc 2005;53(4):695–9. doi: 10.1111/j.1532-5415.2005.53221.x

23. Yesavage JA, Sheikh JI. Geriatric Depression Scale (GDS). Clinical Gerontologist 2008;5(1-2):165–73. doi: 10.1300/J018v05n01_09

24. Buysse DJ, Reynolds CF, 3rd, Monk TH, et al. The Pittsburgh Sleep Quality Index: a new instrument for psychiatric practice and research. Psychiatry Res 1989;28(2):193–213. doi: 10.1016/0165-1781(89)90047-4

25. Mayer JD, Gaschke YN. Brief Mood Introspection Scale. APA PsycTests 1988 doi: 10.1037/t06259-000

26. van Rijsbergen GD, Bockting CL, Berking M, et al. Can a one-item mood scale do the trick? Predicting relapse over 5.5-years in recurrent depression. PLoS One 2012;7(10):e46796. doi: 10.1371/journal.pone.0046796 [published Online First: 20121003]

27. Gershon RC, Wagster MV, Hendrie HC, et al. NIH toolbox for assessment of neurological and behavioral function. Neurology 2013;80(11 Suppl 3):S2–6. doi: 10.1212/WNL.0b013e3182872e5f

28. Fried LP, Tangen CM, Walston J, et al. Frailty in older adults: evidence for a phenotype. J Gerontol A Biol Sci Med Sci 2001;56(3):M146–56. doi: 10.1093/gerona/56.3.m146

29. Zhou L, Bao J, Setiawan IMA, et al. The mHealth App Usability Questionnaire (MAUQ): Development and Validation Study. JMIR Mhealth Uhealth 2019;7(4):e11500. doi: 10.2196/11500 [published Online First: 20190411]

30. Xie Z, Kalun Or C. Acceptance of mHealth by Elderly Adults: A Path Analysis. Proceedings of the Human Factors and Ergonomics Society Annual Meeting 2021;64(1):755–59. doi: 10.1177/1071181320641174

31. Biller AM, Zauner J, Cajochen C, et al. Physiologically-relevant light exposure and light behaviour in Switzerland and Malaysia. J Expo Sci Environ Epidemiol 2025 doi: 10.1038/s41370-025-00825-8 [published Online First: 20251203]

32. Eysenbach G. The law of attrition. J Med Internet Res 2005;7(1):e11. doi: 10.2196/jmir.7.1.e11 [published Online First: 20050331]

33. Milne-Ives M, Lam C, De Cock C, et al. Mobile Apps for Health Behavior Change in Physical Activity, Diet, Drug and Alcohol Use, and Mental Health: Systematic Review. JMIR Mhealth Uhealth 2020;8(3):e17046. doi: 10.2196/17046 [published Online First: 20200318]

34. Hastie T, Tibshirani R. Generalized Additive Models. Statistical Science 1986;1(3) doi: 10.1214/ss/1177013604

35. Zauner J, Guidolin C, Spitschan M. How to Deal With Darkness: Modeling and Visualization of Zero-Inflated Personal Light Exposure Data on a Logarithmic Scale. J Biol Rhythms 2025:7487304251336624. doi: 10.1177/07487304251336624 [published Online First: 20250628]

36. Zauner J, Hartmeyer S, Spitschan M. LightLogR: Reproducible analysis of personal light exposure data. J Open Source Softw 2025;10(107):7601. doi: 10.21105/joss.07601

37. Wickham H, Averick M, Bryan J, et al. Welcome to the Tidyverse. Journal of Open Source Software 2019;4(43) doi: 10.21105/joss.01686

