## Supplementary material for "Protocol for a randomised, double-blind trial of a chronotherapeutic mobile health (mHealth) behaviour change intervention to optimise light exposure among older adults aged ≥60 years in Singapore (LightSPAN)": SPIRIT Checklist

**SPIRIT 2025 checklist of items to address in a randomized trial protocol***

| **Section / Topic** | **No** | **SPIRIT 2025 checklist item description** | **Reported on page no.** |
| --- | --- | --- | --- |
| **Administrative information** | | |  |
| Title and structured summary | 1a | Title stating the trial design, population, and interventions, with identification as a protocol | 1 |
|  | 1b | Structured summary of trial design and methods, including items from the World Health Organization Trial Registration Data Set | 1 |
| Protocol version | 2 | Version date and identifier | 2 |
| Roles and responsibilities | 3a | Names, affiliations, and roles of protocol contributors | 1 |
|  | 3b | Name and contact information for the trial sponsor | 18 |
|  | 3c | Role of trial sponsor and funders in design, conduct, analysis, and reporting of trial; including any authority over these activities | 18 |
|  | 3d | Composition, roles, and responsibilities of the coordinating site, steering committee, endpoint adjudication committee, data management team, and other individuals or groups overseeing the trial, if applicable | 15 |
| **Open science** | | |  |
| Trial registration | 4 | Name of trial registry, identifying number (with URL), and date of registration. If not yet registered, name of intended registry | 2 |
| Protocol and statistical analysis plan | 5 | Where the trial protocol and statistical analysis plan can be accessed | 17 |
| Data sharing | 6 | Where and how the individual de-identified participant data (including data dictionary), statistical code, and any other materials will be accessible | 17 |
| Funding and conflicts of interest | 7a | Sources of funding and other support (e.g., supply of drugs) | 18 |
|  | 7b | Financial and other conflicts of interest for principal investigators and steering committee members | 18 |
| Dissemination policy | 8 | Plans to communicate trial results to participants, healthcare professionals, the public, and other relevant groups (e.g., reporting in trial registry, plain language summary, publication) |  |
| **Introduction** | | |  |
| Background and rationale | 9a | Scientific background and rationale, including summary of relevant studies (published and unpublished) examining benefits and harms for each intervention | 3 |
|  | 9b | Explanation for choice of comparator | 8 |
| Objectives | 10 | Specific objectives related to benefits and harms | 3, 14-15 |
| **Methods: Patient and public involvement, trial design** | | |  |
| Patient and public involvement | 11 | Details of, or plans for, patient or public involvement in the design, conduct, and reporting of the trial | 16-17 |
| Trial design | 12 | Description of trial design including type of trial (e.g., parallel group, crossover), allocation ratio, and framework (e.g., superiority, equivalence, non-inferiority, exploratory) | 4 |
| **Methods: Participants, interventions, and outcomes** | | |  |
| Trial setting | 13 | Settings (e.g., community, hospital) and locations (e.g., countries, sites) where the trial will be conducted | 4 |
| Eligibility criteria | 14a | Eligibility criteria for participants | 4 |
|  | 14b | If applicable, eligibility criteria for sites and for individuals who will deliver the interventions (e.g., surgeons, physiotherapists) | 4 |
| Intervention and comparator | 15a | Intervention and comparator with sufficient details to allow replication including how, when, and by whom they will be administered. If relevant, where additional materials describing the intervention and comparator (e.g., intervention manual) can be accessed | 7-8 |
|  | 15b | Criteria for discontinuing or modifying allocated intervention/comparator for a trial participant (e.g., drug dose change in response to harms, participant request, or improving/worsening disease) | 8 |
|  | 15c | Strategies to improve adherence to intervention/comparator protocols, if applicable, and any procedures for monitoring adherence (e.g., drug tablet return, sessions attended) | 8 |
|  | 15d | Concomitant care that is permitted or prohibited during the trial | 8 |
| Outcomes | 16 | Primary and secondary outcomes, including the specific measurement variable (e.g., systolic blood pressure), analysis metric (e.g., change from baseline, final value, time to event), method of aggregation (e.g., median, proportion), and time point for each outcome | 9-10 |
| Harms | 17 | How harms are defined and will be assessed (e.g., systematically, non-systematically) | 14-15 |
| Participant timeline | 18 | Time schedule of enrollment, interventions (including any run-ins and washouts), assessments, and visits for participants. A schematic diagram is highly recommended (see Figure) | 10-11 |
| Sample size | 19 | How sample size was determined, including all assumptions supporting the sample size calculation | 11-13 |
| Recruitment | 20 | Strategies for achieving adequate participant enrollment to reach target sample size | 12-13 |
| **Methods: Assignment of interventions** | | |  |
| Randomization: |  |  |  |
| Sequence generation | 21a | Who will generate the random allocation sequence and the method used | 5-6 |
|  | 21b | Type of randomization (simple or restricted) and details of any factors for stratification. To reduce predictability of a random sequence, other details of any planned restriction (e.g., blocking) should be provided in a separate document that is unavailable to those who enroll participants or assign interventions | 5-6 |
| Allocation concealment  mechanism | 22 | Mechanism used to implement the random allocation sequence (e.g., central computer/telephone; sequentially numbered, opaque, sealed containers), describing any steps to conceal the sequence until interventions are assigned | 5-6 |
| Implementation | 23 | Whether the personnel who will enroll and those who will assign participants to the interventions will have access to the random allocation sequence | 5-6 |
| Blinding | 24a | Who will be blinded after assignment to interventions (e.g., participants, care providers, outcome assessors, data analysts) | 6-7 |
|  | 24b | If blinded, how blinding will be achieved and description of the similarity of interventions | 6-7 |
|  | 24c | If blinded, circumstances under which unblinding is permissible, and procedure for revealing a participant’s allocated intervention during the trial | 6-7 |
| **Methods: Data collection, management, and analysis** | | |  |
| Data collection methods | 25a | Plans for assessment and collection of trial data, including any related processes to promote data quality (e.g., duplicate measurements, training of assessors) and a description of trial instruments (e.g., questionnaires, laboratory tests) along with their reliability and validity, if known. Reference to where data collection forms can be accessed, if not in the protocol | 11 |
|  | 25b | Plans to promote participant retention and complete follow-up, including list of any outcome data to be collected for participants who discontinue or deviate from intervention protocols | 11 |
| Data management | 26 | Plans for data entry, coding, security, and storage, including any related processes to promote data quality (e.g., double data entry; range checks for data values). Reference to where details of data management procedures can be accessed, if not in the protocol | 11 |
| Statistical methods | 27a | Statistical methods used to compare groups for primary and secondary outcomes, including harms | 13-15 |
|  | 27b | Definition of who will be included in each analysis (e.g., all randomized participants), and in which group | 13 |
|  | 27c | How missing data will be handled in the analysis | 13-14 |
|  | 27d | Methods for any additional analyses (e.g., subgroup and sensitivity analyses) | 14 |
| **Methods: Monitoring** | | |  |
| Data monitoring committee | 28a | Composition of data monitoring committee (DMC); summary of its role and reporting structure; statement of whether it is independent from the sponsor and funder; conflicts of interest and reference to where further details about its charter can be found, if not in the protocol. Alternatively, an explanation of why a DMC is not needed | 15 |
|  | 28b | Explanation of any interim analyses and stopping guidelines, including who will have access to these interim results and make the final decision to terminate the trial | 14 |
| Trial monitoring | 29 | Frequency and procedures for monitoring trial conduct. If there is no monitoring, give explanation | 15 |
| **Ethics** | | |  |
| Research ethics approval | 30 | Plans for seeking research ethics committee/institutional review board approval | 17 |
| Protocol amendments | 31 | Plans for communicating important protocol modifications to relevant parties | 12 |
| Consent or assent | 32a | Who will obtain informed consent or assent from potential trial participants or authorized proxies, and how | 7 |
|  | 32b | Additional consent provisions for collection and use of participant data and biological specimens in ancillary studies, if applicable | 7 |
| Confidentiality | 33 | How personal information about potential and enrolled participants will be collected, shared, and maintained in order to protect confidentiality before, during, and after the trial | 11 |
| Ancillary and post-trial care | 34 | Provisions, if any, for ancillary and post-trial care, and for compensation to those who suffer harm from trial participation | 15 |

*We strongly recommend reading this checklist in conjunction with the SPIRIT 2025 Explanation and Elaboration and the SPIRIT 2025 Expanded Checklist for important clarifications on all the items. We also recommend reading relevant SPIRIT extensions. See [www.consort-spirit.org](http://www.consort-spirit.org)

Citation: Chan A-W, Boutron I, Hopewell S, Moher D, Schulz KF, et al. SPIRIT 2025 statement: updated guideline for protocols of randomised trials. BMJ 2025;389:e081477. <https://dx.doi.org/10.1136/bmj-2024-081477>

© 2025 Chan A-W et al. This is an Open Access article distributed under the terms of the Creative Commons Attribution License (<https://creativecommons.org/licenses/by/4.0/>), which permits unrestricted use, distribution, and reproduction in any medium, provided the original work is properly cited.
