## Supplementary FIgure 1 for "Protocol for a randomised, double-blind trial of a chronotherapeutic mobile health (mHealth) behaviour change intervention to optimise light exposure among older adults aged ≥60 years in Singapore (LightSPAN)"

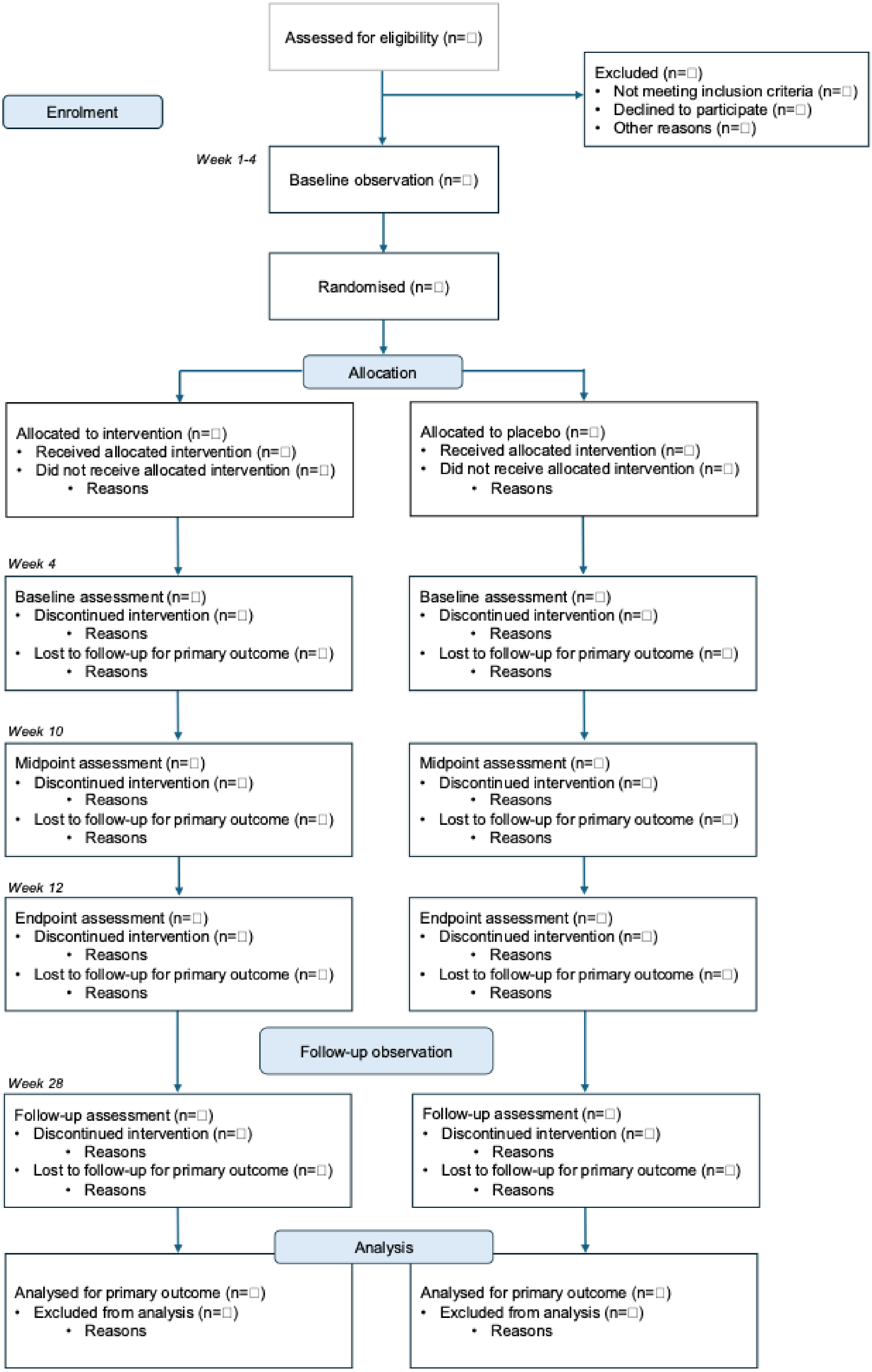


**Supplementary Figure 1.** CONSORT flow diagram illustrating enrolment, randomisation, allocation, assessment time points, follow-up, and analysis for the LightSPAN study protocol.
